# A new design of an adaptive model of infectious diseases based on artificial intelligence approach: monitoring and forecasting of COVID-19 epidemic cases

**DOI:** 10.1101/2020.04.23.20077677

**Authors:** Bachir Nail, Abdelaziz Rabehi, Belkacem Bekhiti, Taha Arbaoui

## Abstract

**Background:** Mathematical infectious disease models available in literature, mostly take in their design that the parameters of basic reproduction number *R*_0_ and interval serial *S*_*I*_ as constant values during tracking the outbreak cases. In this report a new intelligent model called HH-COVID-19 is proposed, with simple design and adaptive parameters.

**Methods:** The parameters *R*_0_ and *S*_*I*_ are adapted by adding three new weighting factors *α, β* and *γ* and two free parameters *σ*_1_ and *σ*_2_ in function of time *t*, thus the HH-COVID-19 become time-variant model. The parameters *R*_0_, *S*_*I*_, *α, β, γ, σ*_1_ and *σ*_2_ are estimated optimally based on a recent algorithm of artificial intelligence (AI), inspired from nature called Harris Hawks Optimizer (HHO), using the data of the confirmed infected cases in Algeria country in the first *t* = 55 days.

**Results:** Parameters estimated optimally: *R*_0_ = 1.341, *S*_*I*_ = 5.991, *α* = 2.987, *β* = 1.566, *γ* = 4.998, *σ*_1_ = *−*0.133 and *σ*_2_ = 0.0324. *R*_0_ starts on 1.341 and ends to 2.677, and *S*_*I*_ starts on 5.991 and ends to 6.692. The estimated results are identically to the actual infected incidence in Algeria, HH-COVID-19 proved its superiority in comparison study. HH-COVID-19 predicts that in 1 May, the infected cases exceed 50 000, during May, to reach quickly the herd immunity stage at beginning of July.

**Conclusion:** HH-COVID-19 can be used for tracking any COVID-19 outbreak cases around the world, just should updating its new parameters to fitting the area to be studied, especially when the population is directly vulnerable to COVID-19 infection.

## 1. Introduction

In Algeria, the first case of COVID-19 was reported on February 25, 2020, when an Italian national tested positive in Ouargla, south of Algeria. On March 1, 2020, two cases were reported in Blida region in the North of Algeria, following their contacts with two Algerian nationals who came from France for holidays. Despite the heightened alert in Algeria to detect and isolated any imported cases of COVID-19, after a few days later, the epidemic of COVID-19 spread to most of Algeria provinces. Until today, 20 April, 2718 confirmed cases with a largest fatality rate in the world 14% (384 deaths) [2].

The proposed mathematical model is developed to fit any outbreak epidemic cases around the world. Where, the following aspects were taken into account in its design:

1. The existed mathematical models not take into account some of the known variables about the disease, and even these variables, such as prevalence rates and mortality ratios, are uncertainties, and clearly change from one region to another.
2. The available mathematical models do not take into account genetic differences (if they have an effect), climate differences (if they have an effect), and previous vaccinations (if they have an impact).
3. The available mathematical models treat the disease as a single mass, while some reports indicate the presence of mutations or changes in the virus, which suggests the presence of different strains, and therefore a different behavior of the disease.

## 2. Methods

The proposed model parameters *R*_0_, and *S*_*I*_ of are designed in such way adapt the uncertainty in the prevalence rates, due to climate differences (if they have an effect), and previous vaccinations (if they have an impact). By adding three small parameters (weighting factors): *α, β* and *γ*. The new parameters 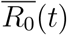 and 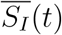 become uncertain (not constant) nonlinear functions in time, this development make the proposed model behavior simulates the reality. The proposed model treats the disease as a multiple masses, by adding two small free parameters *σ*_1_ and *σ*_2_, that can adapt the model toward any mutations or changes in the virus, by the two nonlinear functions *f*_1_(*t*) and *f*_2_(*t*), rrespectively.

The mathematical formulation of the proposed model DZ-COVID-19 is given as follows:

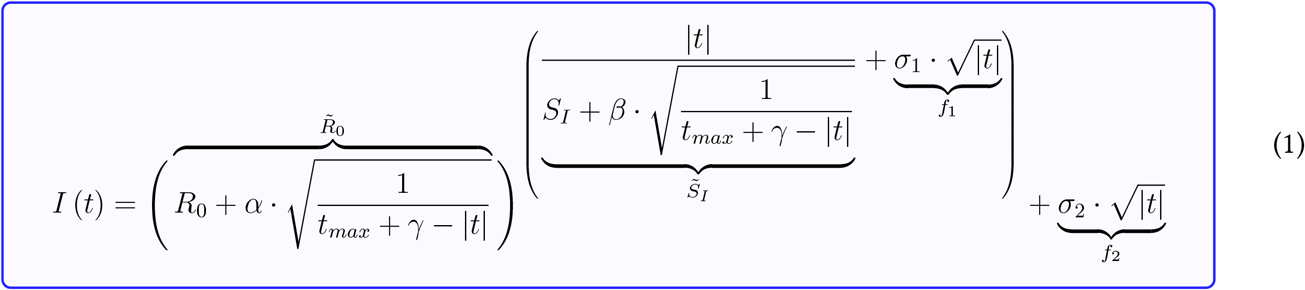

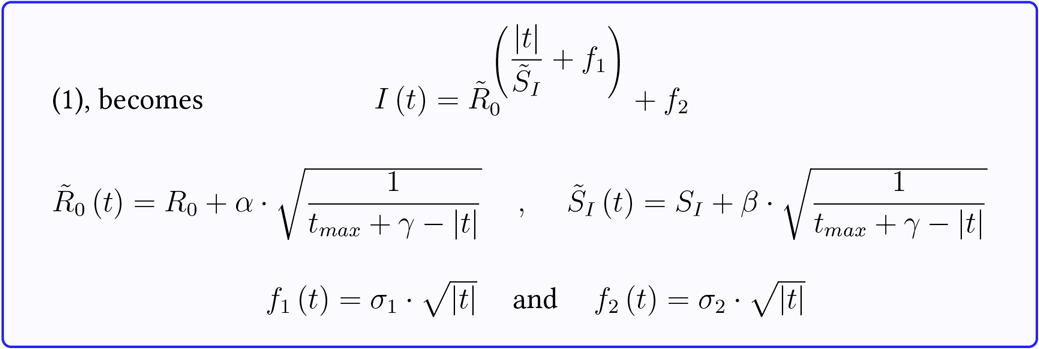

The estimation of DZ-COVID-19 parameters model, presented in (1), first is transformed this objective to convex optimization problem, which solved by the well-known the new optimization algorithm called Harris Hawks Optimizer (HHO). Where, HHO is a metaheuristics algorithm that solve a convex multidimensional optimization problems with constraints [3]. Fig. 1 shows all phases of HHO, Exploration phase, Transition from exploration to exploitation, Exploitation phase, Pseudo-code of HHO, Computational complexity, which are described in detail in [3] and the program codes are in supplementary files.

**Figure 1:**
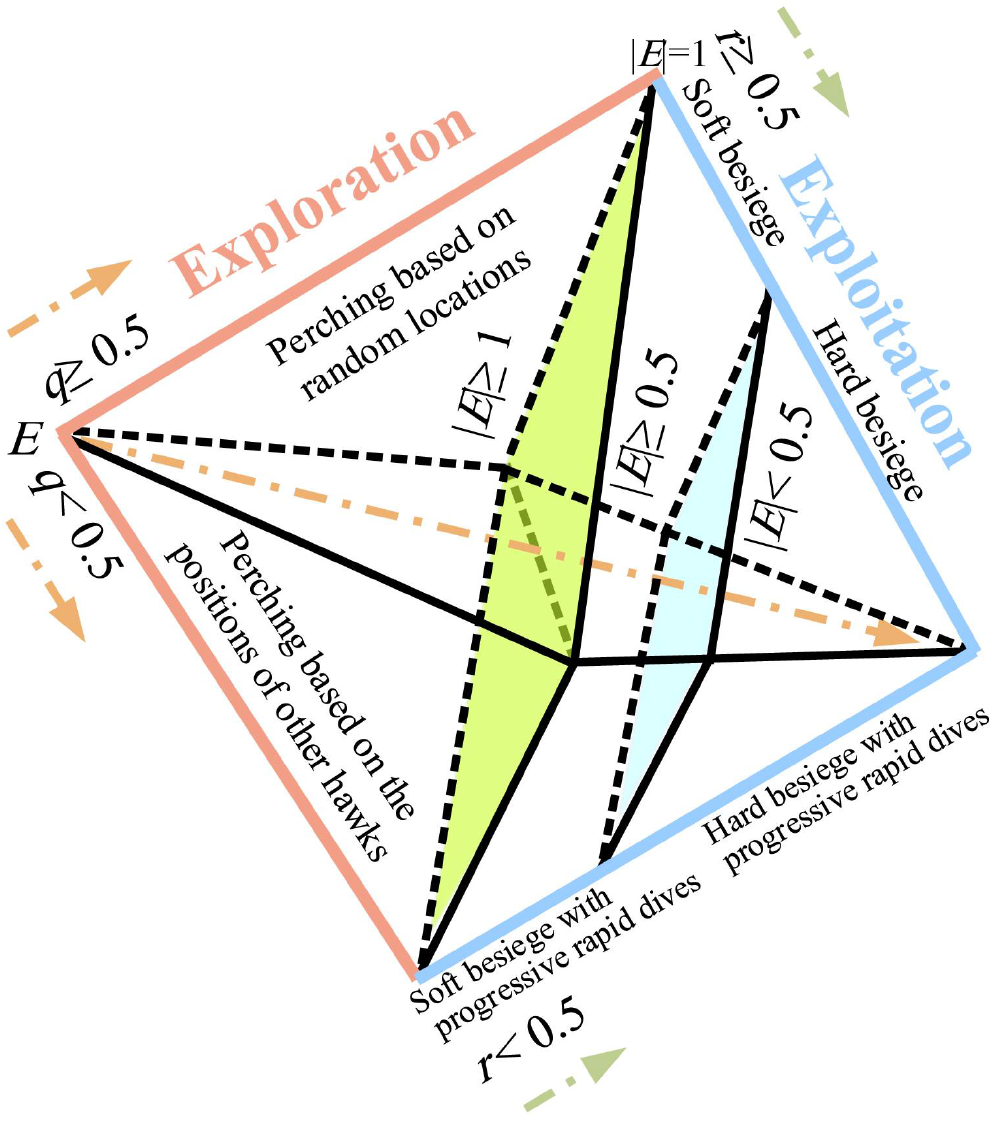
Schematic diagram of different phases of HHO.

The results of this research have been obtained using MATLAB software, (The programs codes are in supplementary files).

The optimization problem of HH-COVID-19 model is given in the form of objective function *f*, which should be minimized to find the optimal parameters of the model HH-COVID-19 *x* = *{R*_0_, *S*_*I*_, *α, β, γ, σ*_1_, *σ*_2_*}*. The objective function *J* is given as follows:

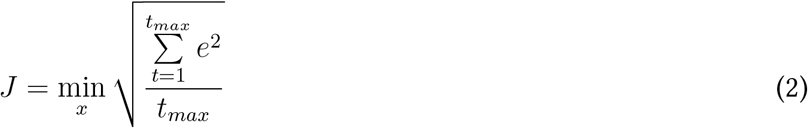

In appendix section you find the parameters setting of HHO used in this estimation.

## 3. Results

## 4. Discussion

Based on obtained results shown in Fig. 4, the proposed model HH-COVID-19 provides an flexibility in tracking, where the estimated HH-COVID-19 incidence cases is coincides with the real data of the infected cases during the finite number of the first 55 days, this feature is lacked in Alg-COVID-19 model [1]. On the other hand, Alg-COVID-19 model leaves the confidence region at 26 March, as shown in Fig. 4. The varying in the parameters of proposed model as shown in Fig. 2, explains that the model adapts any variation in the epidemic behaviour as shown in in Fig. 4 and Fig. 5, this uncertainty in parameters is estimated optimally due to the AI approach using HHO optimizer as shown in Fig. 2.

**Figure 2:**
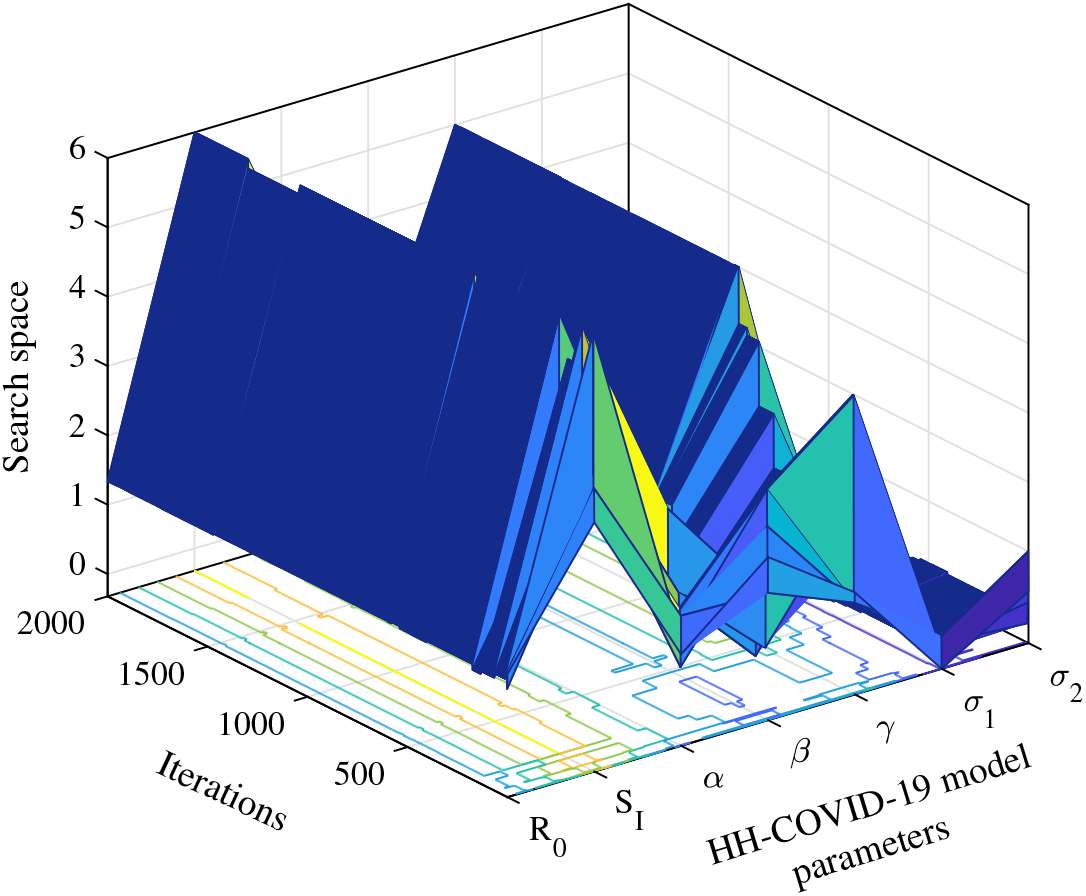
The evolution of HHO to estimating optimal HH-COVID-19 parameters model.

**Figure 3:**
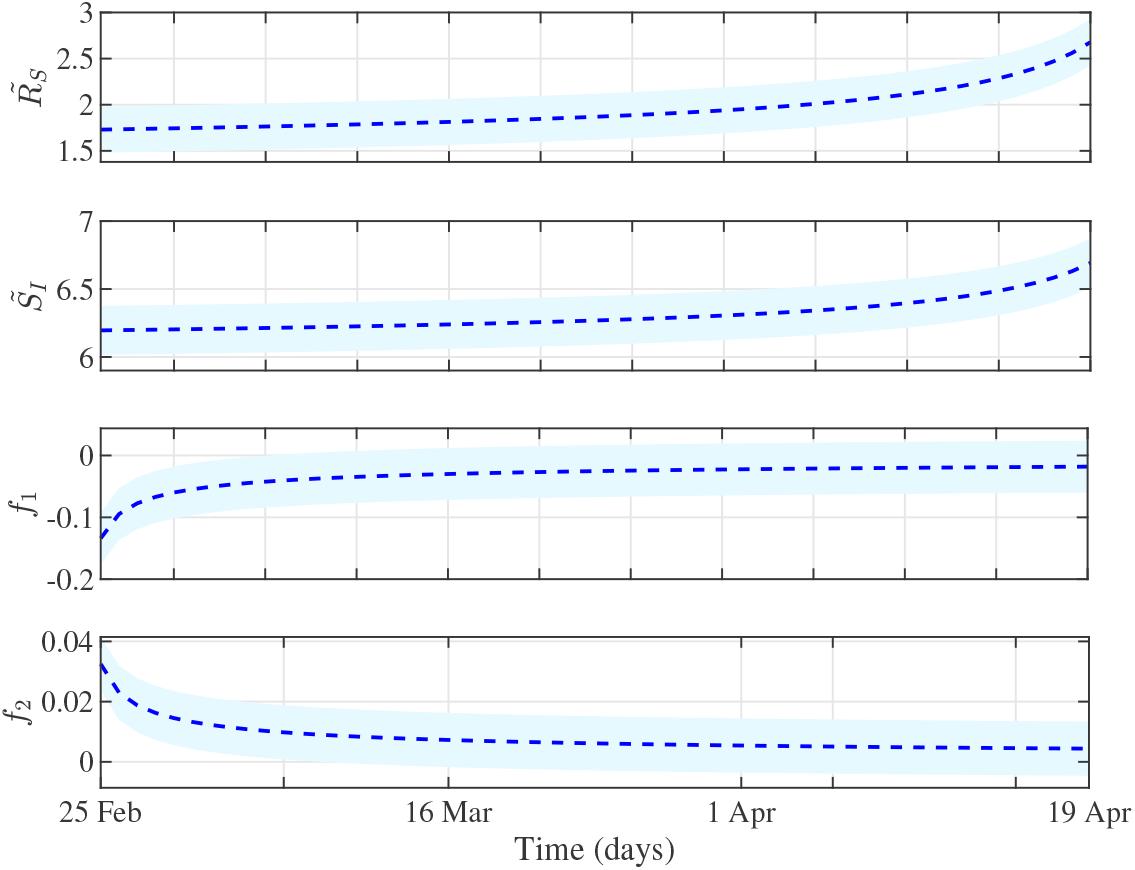
HH-COVID-19 parameters model varying during time.

**Figure 4:**
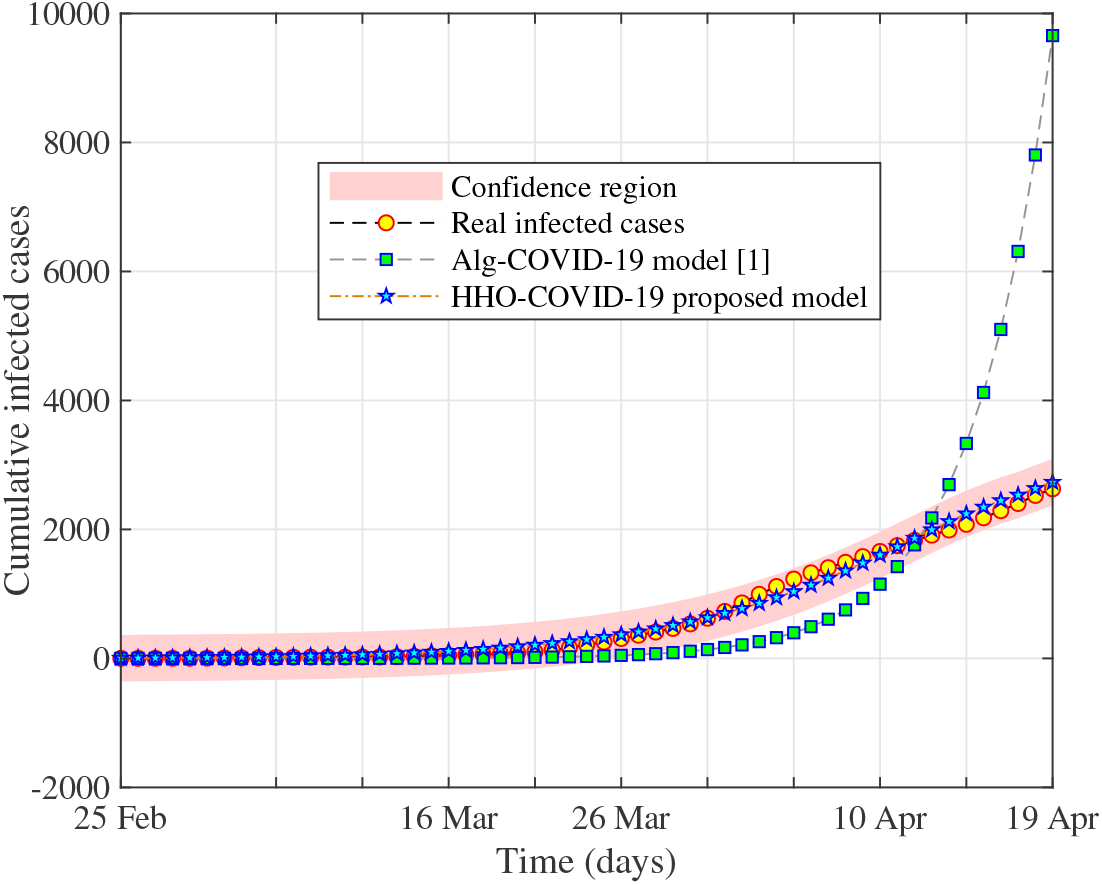
Comparison study between HH-COVID-19 proposed model tracking, Actual infected cases of epidemic during the first 55 days and Alg-COVID-19 model.

**Figure 5:**
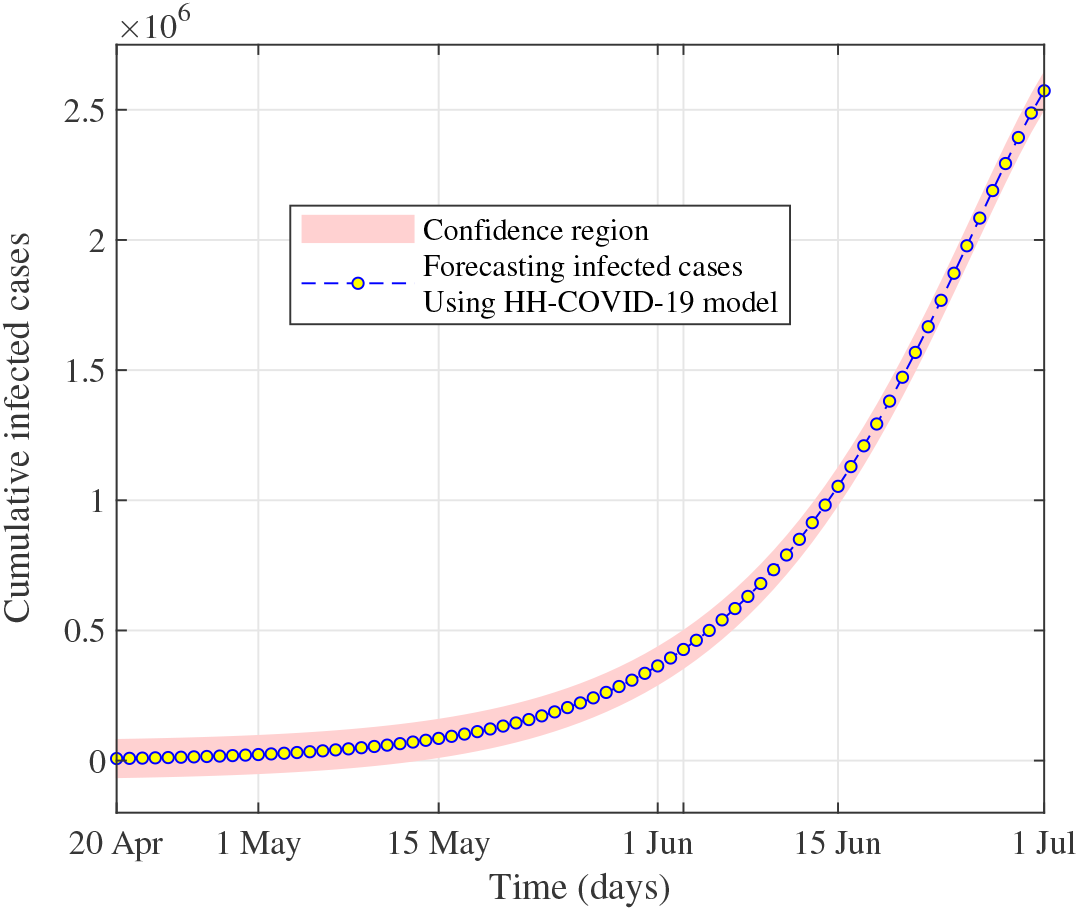
Forecasting the outbreak in the next weeks using HH-COVID-19 model.

According to the very high number of deaths 14% [2], compared to the number of infected cases, with a simple calculation we believe that the real number of incidence cases that should be, if the diagnostic capabilities are available (like as available in developed countries such: USA, China, Italy, France, Spain, … etc) is four times the number declared by the authorities. It means, the number of incidence cases that will be recorded in the next week, is really the cases of this day. Here only to reach the global rate of detection of the disease, which some countries consider a number that does not represent the true number of people infected. If the epidemic still rising at this pace, the parameters of the model is valid, because the most of the population is vulnerable to COVID-19 infection, after 70 days in 1 July, 2020, 2.7 million of people will be infected and we reach to the herd immunity at the end of the same month (July). Except, in the case of *R*_0_ decreases over the next weeks of the epidemic (flatten the curve of Fig. 4). However, this will happen only if the herd immunity rises spontaneously during the epidemic or the decision-makers act on one of the components of *R*_0_ by preventive or curative measures. In this case the parameters of the proposed model HH-COVID-19 should to be updated by estimating a new effective parameters using the new data.

## 5. Data availability

The numbers of confirmed COVID-19 infection cases were obtained based on official daily reports issued by the government institutes (Algerian Ministry of Health 2020), in our country Algeria [2].

## 7. Conclusion

In the best of our knowledge, no one have been proposed this design before, we belive that the proposed model is a reliable, and suitable for tracking epidemics outbreak cases in anywhere around the world, especially COVID-19 epidemic.

## Data Availability

no data availability

## 8. Appendix

**Table 1:** Typical Parameters for HHO

| Parameter | Value |
| --- | --- |
| Search Agents_no | 20 |
| Population size | 7 |
| Lower bound | lb=[1 1 -1 -1 1 -1 -1] |
| Upper bound | ub=[ 4 6 3 3 5 1 1] |
| Iterations | 2000 |

## Reference

[1] Hamidouche M. COVID-19 outbreak in Algeria: A mathematical Model to predict cumulative cases. Bull World Health Organ. E-pub: 25 March 2020. doi:http://dx.doi.org/10.2471/BLT.20.256065

[2] Algerian Ministry of Health. 2020. http://covid19.sante.gov.dz/carte/.

[3] A. A. Heidari, S. Mirjalili, H. Faris, I. Aljarah, M. Mafarja, and H. Chen, “Harris hawks optimization: Algorithm and applications,” Future Generation Computer Systems, vol. 97, pp. 849–872, Aug. 2019.

